# Characterization of Patients Who Return to Hospital Following Discharge from Hospitalization For COVID-19

**DOI:** 10.1101/2020.05.17.20104604

**Authors:** Sulaiman Somani, Felix Richter, Valentin Fuster, Jessica De Freitas, Nidhi Naik, Keith Sigel, The Mount Sinai COVID Informatics Center, Erwin P Bottinger, Matthew A. Levin, Zahi Fayad, Allan C Just, Alexander Charney, Shan Zhao, Benjamin S Glicksberg, Anuradha Lala, Girish N Nadkarni

**Affiliations:** The Hasso Plattner Institute for Digital Health at Mount Sinai, Icahn School of Medicine at Mount Sinai, New York, NY; The Mount Sinai Covid Informatics Center, New York, NY; The Zena and Michael A. Wiener Cardiovascular Institute, Icahn School of Medicine at Mount Sinai, New York, NY; Department of Medicine, Icahn School of Medicine at Mount Sinai; Department of Anesthesiology, Perioperative and Pain Medicine, Icahn School of Medicine at Mount Sinai, New York, NY; The BioMedical Engineering and Imaging Institute, Icahn School of Medicine at Mount Sinai, New York, NY; Department of Environmental Medicine and Public Health, Icahn School of Medicine at Mount Sinai; Department of Genetics and Genomic Sciences, Icahn School of Medicine at Mount Sinai; The Pamela Sklar Division of Psychiatric Genomics, Icahn School of Medicine at Mount Sinai; Icahn Institute for Data Science and Genomic Technology, Icahn School of Medicine at Mount Sinai; Department of Psychiatry, Icahn School of Medicine at Mount Sinai

**Author notes:** co-first authors (Sulaiman Somani, Felix Richter). co-senior authors (Shan Zhao, Benjamin S Glicksberg). co-corresponding authors **Correspondence:** Anuradha Lala, MD, One Gustave Levy Place, Box 1243, New York, NY 10029, Girish N Nadkarni, MD, MPH, One Gustave Levy Place, Box 1243, New York, NY 10029.

## Abstract

**Background:** Data on patients with coronavirus disease 2019 (COVID-19) who return to hospital after discharge are scarce. Characterization of these patients may inform post-hospitalization care.

**Methods and Findings:** Retrospective cohort study of patients with confirmed SARS-CoV-2 discharged alive from five hospitals in New York City with index hospitalization between February 27^th^-April 12^th^, 2020, with follow-up of ≥14 days. Significance was defined as *P*<0.05 after multiplying *P* by 125 study-wide comparisons. Of 2,864 discharged patients, 103 (3.6%) returned for emergency care after a median of 4.5 days, with 56 requiring inpatient readmission. The most common reason for return was respiratory distress (50%). Compared to patients who did not return, among those who returned there was a higher proportion of COPD (6.8% vs 2.9%) and hypertension (36% vs 22.1%). Patients who returned also had a shorter median length of stay (LOS) during index hospitalization (4.5 [2.9,9.1] vs. 6.7 [3.5, 11.5] days; *P_adjusted_*=0.006), and were less likely to have required intensive care on index hospitalization (5.8% vs 19%; *P_adjusted_*=0.001). A trend towards association between absence of in-hospital anticoagulation on index admission and return to hospital was also observed (20.9% vs 30.9%, *P_adjusted_*=0.064). On readmission, rates of intensive care and death were 5.8% and 3.6%, respectively.

**Conclusions:** Return to hospital after admission for COVID-19 was infrequent within 14 days of discharge. The most common cause for return was respiratory distress. Patients who returned had higher proportion of COPD and hypertension with shorter LOS on index hospitalization, and a trend towards lower rates of in-hospital treatment-dose anticoagulation. Future studies should focus on whether these comorbid conditions, longer LOS and anticoagulation are associated with reduced readmissions.

## INTRODUCTION

Despite four months after the first cluster of cases were reported in Wuhan, China, coronavirus disease 2019 (COVID-19) persists as a major cause of morbidity and mortality worldwide.^1^ Healthcare facilities remain stretched beyond capacity, further burdened by readmissions in a subset of the hospitalized population. As the evolution of COVID-19 remains uncertain, characterization of the clinical course of patients who represent after discharge is important with potential to inform discharge planning and post-discharge care.^2^

## METHODS

### Study Population and Data Collection

We included patients at least 18 years old with laboratory-confirmed SARS-CoV-2 infection who were admitted and subsequently discharged alive from five MSHS hospitals. Data were collected from electronic health records (EHR). Patients were excluded if discharge occurred after April 12, 2020 to ensure all patients had at least a 14-day observation for possible readmission. A COVID-related hospital admission included any inpatient encounter with detected SARS-CoV-2 from an order within the first 48 hours of the encounter, including time spent in the emergency department, or within 48-hours of an antecedent lab diagnosis-only visit to an outpatient setting. Given the vast amount of information contained in the EHR, variables were chosen in this analysis based on a combination of both their prevalence per patient across the dataset, relevance to COVID based on previous literature and empirical evidence, and appropriate goals of this study. This variable selection, culling, and cleanup was performed through exhaustive manual inspection by clinicians. Labs with >80% missingness were subsequently excluded, resulting in an a priori set of 125 variables for comparison of the primary endpoint. Of these 125 variables (all of which are accounted for through multiple hypothesis testing), 66 are shown in Table 1 and Supplemental Table 1 to highlight pertinent positive and negative findings most salient to results interpretation. Patients were excluded if they were still hospitalized, discharged < 14 days prior to the data freeze on April 26th, 2020, returned <12 hours after discharge, or died during the index admission. Variables collected included demographics, vitals and laboratory measurements on time of discharge from index hospitalization, disease diagnoses, comorbidities, procedures during hospitalization (including intubation and non-invasive O_2_ oxygen support), ICU course, and outcomes (death or hospital discharge). Comorbidities were extracted using International Classification of Disease (ICD) 9/10 billing codes for atrial fibrillation (AF), asthma, coronary artery disease (CAD), cancer, chronic kidney disease (CKD), chronic obstructive pulmonary disease (COPD), diabetes (DM), heart failure (HF), and hypertension (HTN). History and physical exam findings were extracted from the initial note with a natural language processing (NLP) algorithm cross-validated against findings extracted manually by medical students. Chest X-rays were screened for infiltrates from the corresponding report by the radiologist using an NLP algorithm similarly cross-validated against findings extracted manually by medical students. In-hospital anticoagulants included treatment-dose oral anticoagulation (warfarin, dabigatran, rivaroxaban, apixaban, edoxaban), treatment dose low molecular weight heparin (bemiparin, certoparin, dalteparin, enoxaparin, nadroparin, parnaparin, reviparin, tinzaparin), and intravenous heparin. The Mount Sinai Institutional Review Board approved this research under a broad research protocol for patient-level data analysis.

### Statistical Analysis

Medians and interquartile ranges were used for continuous data. Univariate statistical significance was identified with the non-parametric Kruskal-Wallis one-way ANOVA or chi-squared tests at Bonferroni-adjusted *P*<0.05 (multiplying *P* by 125 study-wide tests). Due to variable correlations, Bonferroni correction ensured a conservative interpretation. P-values were adjusted separately within each sub-group, considered distinct hypothesis spaces. Wilcoxon signed-rank tests compared pairwise differences in labs between discharge and return. Time to hospital return was compared with cumulative density function plots. Analyses were performed using R (R Foundation) and Python (Python Software Foundation).

## RESULTS

### Patient and Index Hospitalization Associations with Return to Hospital

Of 2,864 patients, 103 (3.6%) returned to one of five hospitals within 14 days, with 56 (54.4%) readmitted and the remaining 44 (45.6%) discharged from the emergency department (ED) **(Table 1)**. There were no between-group differences in age, sex, or race/ethnicity; however, patients who returned to the hospital had lower BMI (26.1 vs. 28.0 kg/m^2^, *P_adjusted_*<0.001). Chronic obstructive pulmonary disease (COPD) (6.8% vs 2.9%, *P_adjusted_*=0.035) and hypertension (36% vs 22.1%, *P_adjusted_*=0.003) were more common in those who returned, though other comorbidities such as chronic kidney disease, asthma, coronary artery disease, and diabetes mellitus were not. Though not statistically significant, a clinically significant in a lower frequency of anticoagulation use among patients who re-presented to the hospital when compared to those who did not re-present to the hospital (20.9 vs 30.9%, *P_adjusted_*=0.06) was observed.

**Table 1:** Clinical Characteristics of All Patients Re-Presenting to the Hospital.

| | No Return to Hospital | Return to Hospital | Univariate $P_{adjusted}^*$ | N Missing |
| --- | --- | --- | --- | --- |
| N (%) | 2761 (96.4) | 103 (3.6) |  |  |
| Age - median (IQR) | 65.9 [54.5,77.0] | 66.1 [53.7,75.1] | 0.538 | 0 |
| Sex- no. (%) |  |  |  |  |
| Female | 1163 (42.1) | 38 (36.9) | 0.340 | 0 |
| Male | 1598 (57.9) | 65 (63.1) |  |  |
| Race/Ethnicity (%) |  |  |  |  |
| Asian | 110 (4.0) | 4 (3.9) | 0.434 | 0 |
| Black/African American | 776 (28.1) | 37 (35.9) |  |  |
| Hispanic/Latino | 754 (27.3) | 27 (26.2) |  |  |
| Other | 376 (13.6) | 10 (9.7) |  |  |
| Unknown | 87 (3.2) | 1 (1.0) |  |  |
| White | 658 (23.8) | 24 (23.3) |  |  |
| BMI - median (IQR) | 28.0 [24.7,32.6] | 26.1 [23.1,30.6] | 0.005 | 330 |
| Comorbidities - no. (%) |  |  |  |  |
| Atrial fibrillation | 106 (3.8) | 4 (3.9) | 1.000 | 0 |
| Asthma | 104 (3.8) | 6 (5.8) | 0.288 | 0 |
| CAD | 220 (8.0) | 12 (11.7) | 0.246 | 0 |
| Cancer | 104 (3.8) | 7 (6.8) | 0.118 | 0 |
| CKD | 128 (4.6) | 8 (7.8) | 0.152 | 0 |
| COPD | 80 (2.9) | 7 (6.8) | 0.035 | 0 |
| Diabetes | 420 (15.2) | 19 (18.4) | 0.450 | 0 |
| Heart failure | 159 (5.8) | 8 (7.8) | 0.522 | 0 |
| Hypertension | 610 (22.1) | 36 (35.0) | 0.003 | 0 |
| Stroke | 83 (3.0) | 1 (1.0) | 0.369 | 0 |
| Liver disease | 67 (2.4) | 3 (2.9) | 0.739 | 0 |
| Non-invasive O2 support - no. (%) | 1347 (48.8) | 43 (41.7) | 0.193 | 0 |
| Any Anticoagulant - no. (%) | 737 (30.9) | 18 (20.9) | 0.064 | 392 |
| Last vitals - median (IQR) |  |  |  |  |
| Systolic Blood Pressure, mm Hg | 119.0 [107.0,133.0] | 126.0 [115.5,135.5] | 0.002 | 1 |
| Diastolic Blood Pressure, mm Hg | 70.0 [61.0,77.0] | 71.0 [64.0,79.5] | 0.175 | 1 |
| Temperature, F | 98.0 [97.4,98.6] | 98.1 [97.5,98.7] | 0.626 | 1 |
| Heart Rate, bpm | 85.0 [74.0,96.2] | 84.0 [74.0,94.0] | 0.269 | 1 |
| Pulse oximetry, % | 95.0 [93.0,97.0] | 96.0 [94.0,98.0] | 0.022 | 6 |
| Respiration Rate, breaths/minute | 18.0 [18.0,20.0] | 18.0 [18.0,19.0] | 0.040 | 1 |
| **Last EKG QTc - median (IQR)** | 450.0 [431.0,474.0] | 447.0 [430.0,475.0] | 0.888 | 348 |
| **Metabolic markers** |  |  |  |  |
| Sodium, MeQ/L | 139.0 [137.0,142.0] | 138.0 [135.0,141.0] | <0.001 | 60 |
| Potassium, MeQ/L | 4.4 [4.0,4.8] | 4.2 [3.8,4.6] | 0.001 | 53 |
| Blood Urea Nitrogen, mg/dL | 18.0 [12.0,37.0] | 18.0 [11.0,30.0] | 0.362 | 51 |
| Creatinine, mg/dL | 0.9 [0.7,1.6] | 0.9 [0.7,1.5] | 0.795 | 41 |
| Glucose, mg/dL | 106.0 [89.0,151.0] | 99.0 [85.0,121.2] | 0.013 | 48 |
| Calcium, mEq/L | 8.4 [7.9,8.8] | 8.3 [7.8,8.8] | 0.803 | 49 |
| **Hematological markers** |  |  |  |  |
| White Blood Cells, 10<sup>3</sup>/μL | 7.7 [5.6,10.7] | 6.3 [4.9,8.7] | 0.001 | 8 |
| Hemoglobin, mEq/L | 12.0 [10.1,13.4] | 11.7 [10.0,13.4] | 0.892 | 16 |
| Neutrophil Percentage | 73.1 [62.7,82.7] | 70.4 [63.1,79.8] | 0.101 | 20 |
| Lymphocyte Percentage | 16.2 [9.5,24.0] | 16.7 [10.7,24.2] | 0.219 | 20 |
| Platelets, no. | 262.0 [186.0,373.2] | 231.0 [180.5,337.5] | 0.091 | 9 |
| **Liver function** |  |  |  |  |
| Total Bilirubin, mg/dL | 0.6 [0.4,0.8] | 0.5 [0.4,0.7] | 0.359 | 102 |
| Gamma-Glutamyl Transpeptidase, units/L | 48.0 [30.0,100.0] | 35.0 [21.0,134.8] | 0.317 | 2225 |
| Albumin, g/dL | 2.8 [2.4,3.2] | 3.0 [2.6,3.4] | <0.001 | 102 |
| **Coagulation markers** |  |  |  |  |
| International Normalized Ratio | 1.1 [1.1,1.3] | 1.1 [1.0,1.2] | 0.162 | 1179 |
| **Inflammatory markers** |  |  |  |  |
| C Reactive Protein, mg/L | 66.0 [24.2,143.0] | 70.9 [31.3,143.6] | 0.696 | 478 |
| D-Dimer, ng/mL | 1.6 [0.8,3.2] | 1.8 [0.8,2.9] | 0.696 | 719 |
| Erythrocyte Sedimentation Rate, mm/hour | 64.0 [37.8,90.0] | 72.0 [44.5,89.8] | 0.652 | 1632 |
| Ferritin, ng/mL | 749.5 [380.0,1607.2] | 590.0 [305.0,1238.5] | 0.086 | 506 |
| Fibrinogen | 612.0 [494.0,742.5] | 621.0 [514.0,747.0] | 0.775 | 1628 |
| Interleukin- 1β | 0.4 [0.3,0.7] | 0.5 [0.3,0.8] | 0.581 | 2234 |
| Interleukin- 6 | 56.8 [25.1,124.0] | 45.4 [26.3,77.9] | 0.128 | 1337 |
| Lactate Dehydrogenase, units/L | 394.0 [304.0,512.0] | 316.0 [255.0,437.0] | <0.001 | 566 |
| Procalcitonin, ng/mL | 0.2 [0.1,0.6] | 0.2 [0.1,0.7] | 0.841 | 681 |
| **Cardiac markers** |  |  |  |  |
| B-Type Natriuretic Peptide, pg/mL | 67.5 [26.0,212.7] | 92.1 [32.3,242.1] | 0.192 | 1362 |
| Troponin, ng/mL | 0.0 [0.0,0.1] | 0.0 [0.0,0.1] | 0.970 | 414 |
| **ICU admission - no. (%)** | 524 (19.0) | 6 (5.8) | 0.001 | 0 |
| **ICU Length of stay, days - median (IQR)** | 4.6 [1.4,9.9] | 2.4 [1.1,5.0] | 0.376 | 0 |
| **Intubated - no (%)** | 293 (10.6) | 1 (1.0) | 0.003 | 0 |
| **Length of stay, days - median (IQR)** | 6.7 [3.5,11.5] | 4.7 [2.9,9.1] | 0.006 | 0 |

\**P*-values were multiplied by 125 to conservatively account for all 125 hypothesis tests (Bonferroni correction), of which 66 are shown based on relevance to study design and outcomes.

Across the entire subset of individuals that re-presented to the hospital, the median time to return was 4.5 days (**Fig 1**). Among patients who returned to the hospital, we observed a shorter median index hospitalization LOS (4.7 days; 95% confidence interval (CI) 2.9-9.1), compared to patients who did not return (6.7 days, CI 3.5-11.7, *P_adjusted_*=0.006) (**S1 Table**). This trend was consistent in subgroups restricted to patients who were intubated (*P_adjusted_*=0.006) or admitted to the ICU (*P_adjusted_*=0.003) during index admission.

**Fig 1.**
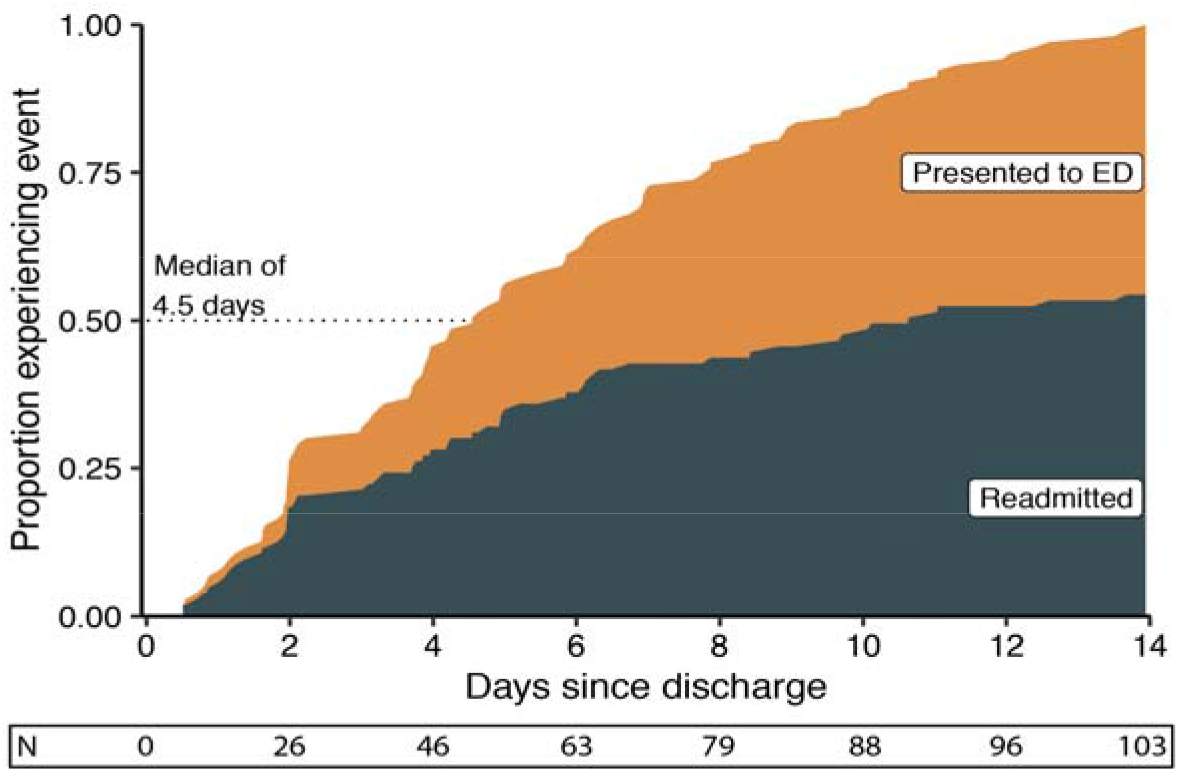
Time to return to hospital since discharge for Covid-19-related hospitalization. Cumulative density function of time to return to the ED or readmission illustrates the proportion of patients (N=103) from each return visit type contributing to the total. For every time point, the proportion of readmitted patients (N=56) is higher than those who are only seen in the ED and then discharged (N=47).

Certain index hospitalization laboratory values on discharge differed among patients who returned vs. those who did not (**Table 1**). These included minor differences in components of the basic metabolic panel, including sodium, potassium, and glucose. Higher white blood cell (WBC) counts, neutrophil percentages, albumin, and INR, as well as lower lymphocyte percentages and LDH, were observed on discharge among patients who returned. However, no major differences were found in other recorded inflammatory markers, including C-reactive protein, erythrocyte sedimentation rate (ESR), fibrinogen, ferritin, procalcitonin, or troponin.

### Reasons for and Outcomes on Return

The most common reason for return to hospital was respiratory distress (50%). Other reasons included chest pain (6%), other pain (6%), altered mental status (5%), falls (5%), and soft tissue infections (5%) (**Fig 2**), consistent in subgroups readmitted and discharged from the ED. In comparison to vitals taken at discharge from the index hospitalization, pulse (96 vs. 82 bpm) and respiratory rate (19 vs. 18 breaths/min) were higher on return to hospital. Returning patients similarly had higher WBC counts (9.2×10^3^ vs. 7.0×10^3^ cells), and lower lymphocyte percentages (12.1% vs 14.8%) compared to discharge (**S2 Table**). On readmission, six (10.7%) patients required ICU-level care. Among all patients that were readmitted upon return, 51 were (91.1%) successfully discharged, three (5.4%) died, and two (3.6%) remained hospitalized.

**Fig 2:**
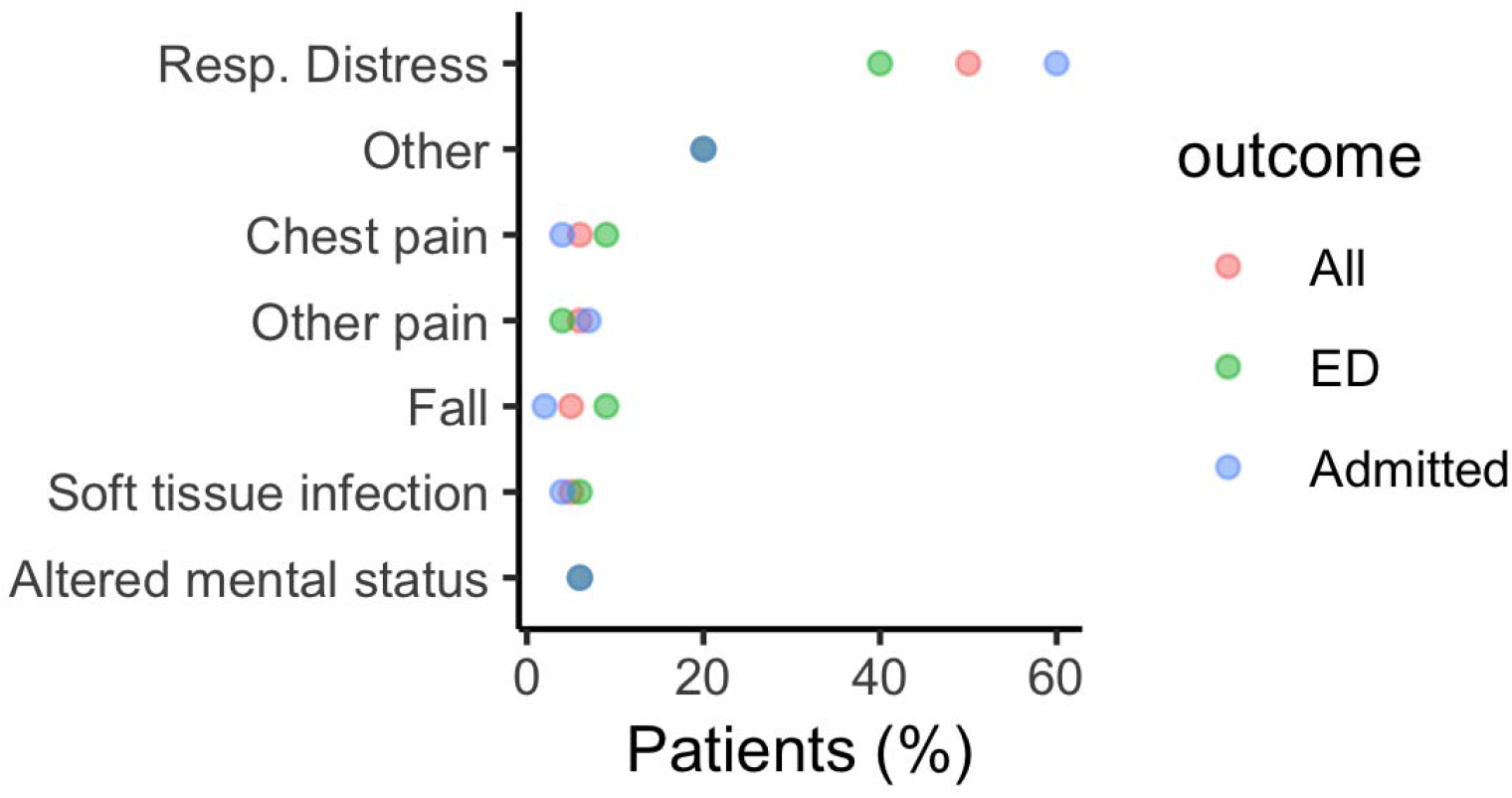
Reasons for return to hospital in 103 patients. Among patients who returned to hospital within the 14-day event horizon (N=103), the majority presented with respiratory distress as the primary reason.

## DISCUSSION

To the best of our knowledge, this is the first report characterizing early return to hospital following discharge in COVID-19 patients. Though the percentage of patients returning to hospital within 14 days was small, we made several important observations. First, patients who returned generally presented within 5 days of discharge, and of these, only half required hospital readmission. Age, sex, and race/ethnicity were not different in patients returning to the hospital compared to those who did not return, however there was a higher prevalence of hypertension and COPD. Second, the most common cause for returning to hospital was respiratory distress accounting for half of all symptoms on return. Finally, returning patients had shorter LOS on index hospitalization with notably a lower frequency of ICU care and inpatient anticoagulation. These findings may offer implications for the post-discharge care of patients hospitalized with COVID-19, and if verified, could inform resource allocation.^3,4^

As the number of patients hospitalized with COVID-19 continues to grow, understanding the frequency and nature of readmissions is essential. It was interesting to note that patients returned to hospital relatively early after discharge at a median of 4.5 days. While the lack of association between age and return to hospital was surprising, the finding that COPD was more common among returning patients was consistent with the observation that respiratory distress was the most common cause for return.

Additionally, the lower BMI observed in those who returned has implications for the role of frailty. Specifically, lower BMI could serve as a marker for and has been associated worse health outcomes in other disease states. Further work is needed to understand the association between BMI and outcomes in COVID-19.

We also note that patients who returned to the hospital had lower LOS on index hospitalization. Despite administrative prioritization to reduce LOS and readmissions, studies across a variety of disease states have suggested these two outcomes may be inversely related.^5–7^ Reduced LOS during the COVID-19 pandemic has also been emphasized for the purposes of preserving resources and limiting exposure. Further, we observed returning patients were less likely to have required ICU stay during the index hospitalization. ICU stay serves as a marker of illness severity and thereby may caution practitioners to ensure clinical stability prior to discharge.^8^ Whether continued in-hospital observation translating into longer LOS for improvement in respiratory status impacts readmission status and exposure risk warrants further study.^9, 10^

Finally, we observed a trend towards the association between less frequent anticoagulation use and return to hospital. There is increasing speculation that COVID-19 is a pro-thrombotic disease ^9,10^ and as such, the impact of anticoagulation on readmission and other outcomes are of special interest.

Several limitations of this study warrant mention. First, small sample sizes restricted statistical power and prevented multivariable analysis to adequately control for non-normal distributions and feature collinearities.^11,12^ Larger sample sizes may allow the development of such multivariable models to address potentially confounding factors and are actively being pursued.^13^ Second, although the MSHS reflects a large and diverse cohort, clinical management varies across hospitals and continues to evolve. Additionally, many of these discharges were in the earlier period of the COVID-19 pandemic when hospital capacity was strained. Thus, generalizability may be limited due to a possible temporal bias, necessitating extension of current studies to longer time periods. Third, readmission over a 30-day horizon may permit comparative analyses with readmission rates for other diseases to inform impact on systems-level operations. Finally, the number of readmissions may have been underreported due to presentation to hospitals outside of the Mount Sinai Health system that could not be tracked via EHR.

## CONCLUSIONS

Among patients discharged following admission for COVID-19, the rate of return to hospital was relatively low, with only half requiring readmission. Patients who returned had respiratory distress and were more likely to have a history of COPD and HTN. They importantly also had a shorter length of stay and lower frequency of anticoagulation use during their index hospitalization. Future work should be directed to understand determinants of safe discharge and appropriate in-hospital treatment to prevent readmission following hospitalization for COVID-19.

## Data Availability

Individual level data are not available for this study. Aggregate data are available in the main and supplemental tables. Please contact the corresponding author for additional data.

## References

1. Khachfe HH, Chahrour M, Sammouri J, Salhab H, Makki BE, Fares M. An Epidemiological Study on COVID-19: A Rapidly Spreading Disease. Cureus. 2020;12(3):e7313.

2. Richardson S, Hirsch JS, Narasimhan M, et al. Presenting Characteristics, Comorbidities, and Outcomes Among 5700 Patients Hospitalized With COVID-19 in the New York City Area. JAMA. April 2020. doi:10.1001/jama.2020.6775

3. Bueno H, Ross JS, Wang Y, et al. Trends in length of stay and short-term outcomes among Medicare patients hospitalized for heart failure, 1993–2006. JAMA. 2010;303(21):2141–2147.

4. Jencks SF, Williams MV, Coleman EA. Rehospitalizations among patients in the Medicare fee-for-service program. N Engl J Med. 2009;360(14):1418–1428.

5. Eapen ZJ, Reed SD, Li Y, et al. Do countries or hospitals with longer hospital stays for acute heart failure have lower readmission rates?: Findings from ASCEND-HF. Circ Heart Fail. 2013;6(4):727–732.

6. Damrauer SM, Gaffey AC, DeBord Smith A, Fairman RM, Nguyen LL. Comparison of risk factors for length of stay and readmission following lower extremity bypass surgery. J Vasc Surg. 2015;62(5):1192–1200.e1.

7. Sud M, Yu B, Wijeysundera HC, et al. Associations Between Short or Long Length of Stay and 30-Day Readmission and Mortality in Hospitalized Patients With Heart Failure. JACC: Heart Failure. 2017;5(8):578–588. doi:10.1016/j.jchf.2017.03.012

8. Chalmers JD. ICU admission and severity assessment in community-acquired pneumonia. Crit Care. 2009;13(3):156.

9. Ruan Q, Yang K, Wang W, Jiang L, Song J. Correction to: Clinical predictors of mortality due to COVID-19 based on an analysis of data of 150 patients from Wuhan, China. Intensive Care Med. April 2020. doi:10.1007/s00134-020-06028-z

10. Mehta P, McAuley DF, Brown M, et al. COVID-19: consider cytokine storm syndromes and immunosuppression. Lancet. 2020;395(10229):1033–1034.

11. Song Y, Morency L, Davis R. Distribution-sensitive learning for imbalanced datasets. In: 2013 10th IEEE International Conference and Workshops on Automatic Face and Gesture Recognition (FG).; 2013: 1–6.

12. Sharma BR, Kaur D, Manju A. Review on Data Mining: Its Challenges, Issues and Applications. International Journal of Current Engineering and Technology. 2013;3(2).

13. Biau DJ, Kernéis S, Porcher R. Statistics in brief: the importance of sample size in the planning and interpretation of medical research. Clin Orthop Relat Res. 2008;466(9):2282–2288.

